# A Novel Methodology to Recalibrate Pathogenic Range of SCA36 Repeat Expansions for PGT-M

**DOI:** 10.1101/2024.08.11.24311662

**Authors:** Fulin Liu, Wen Huang, Ling Liao, Jiyun Yang

## Abstract

**Background:** Spinocerebellar ataxia-36 (SCA36) is an inherited neurodegenerative disorder caused by the heterozygous expansion of an intronic GGCCTG hexanucleotide repeat in the NOP56 gene on chromosome 20p13. Unaffected individuals typically carry 3 to 14 repeats, whereas affected individuals carry 650 to 2,500. However, based on a single study, this pathogenic range was conservatively established, limiting its extended clinical applicability such as preimplantation genetic testing (PGT). In this study, we propose a novel methodology to recalibrate the pathogenic range of SCA36 repeat expansion.

**Methods:** We conducted a comprehensive literature review and collected examination data from 2012 onward. We used the gamma distribution to describe the data distribution and applied Bayesian methods to update the prior distribution with data from recent publications. Based on the recalibrated distribution, the 95% confidence interval (CI) was used to determine the new lower boundary of the pathogenic range. A pedigree was collected to validate the proposal with long-read sequencing (LRS) applied to detect the high GC content and long length of repeat expansions.

**Results:** Our results, based on 2 studies, indicate that the data distribution is well-described by gamma distribution. The prior, likelihood and posterior distributions within the 95% CI for the integrated research of SCA36 pathogenic repeat expansions were [446, +∞), [124, +∞), and [484, +∞), respectively. These recalibrated pathogenic ranges were validated by an authentic case: a proband diagnosed with SCA36 carrying 418 repeats and her daughter with 499 repeats, under the detection of LRS.

**Conclusions:** Therefore, we proposed a novel methodology that integrates updated data, 95% CI using Bayesian methods and LRS for accurate detection of repeat expansions of dynamic mutations to present an up-to-date pathogenic range of SCA36, as well as other similar diseases.

## Background

The human genome contains a substantial number of tandem repeats (TRs). TRs exhibit a high mutation rate within the genome due to their repetitive nature, resulting in enormous polymorphism and multiallelism. The variability of TRs in the length of the repeat sequences occurs in both non-coding and coding regions of the genome [1]. A certain range of TR expansions beyond a critical threshold can lead to disease onset and may increase or decrease across generations, which is the foundation of dynamic mutations [2]. The intergenerational expansion of dynamic mutations is a well-documented phenomenon that underlies the pathogenesis of at least 50 known disorders [3].

Clinical diagnosis of dynamic mutations necessitates not only the observation of characteristic clinical manifestations but also the application of auxiliary diagnostic tests, with genetic testing being paramount. Diagnosis depends on identifying the number of dynamic mutations within a defined pathogenic range. When the mutation count falls within the defined range, a clinical diagnosis can be made; otherwise, a negative result is inferred. However, there exists an uncertain range of clinical significance beyond the confirmatory and exclusion ranges. Currently, the evaluation of the uncertain range primarily relies on clinical diagnosis. If clinical diagnosis is definitive, the uncertain range is considered positive; if clinical phenotypes cannot be correlated, it is deemed negative. Nonetheless, clinical practice often encounters cases where phenotypes are ambiguous, and genetic testing results fall within the uncertain range. This poses ethical challenges, particularly for preimplantation genetic testing (PGT) candidates, where ethics committees may not approve the subsequent procedure according to the guidelines [4–6].

PGT, especially PGT for monogenic disorders (PGT-M), aims to enable pregnancies unaffected by specific genetic traits carried by one or both parents. PGT-M offers couples with genetic disorders the opportunity to have their offspring and avoid the birth of defective fetuses, yet its ethical standards are stringently ruled. This stringency stems from the current limitations in basic medical research to fully elucidate the relationship between genetic variants and clinical phenotypes, leading to poor gene-disease mapping. Thus, strict inclusion criteria are employed to minimize off-target resulting in defective fetuses. Namely, PGT-M can be only administered in a certain pathogenic range but not the uncertain range of repeat expansions, which poses a significant challenge to the uncertainty. Such stringent criteria may result in too many considerations to perform the PGT-M. Therefore, this study proposes a novel but reasonable methodology to recalibrate the defined pathogenic range of repeat expansions so that more patients within the uncertain range win steady support for the PGT-M.

Taking spinocerebellar ataxia type 36 (SCA36) as an example, SCA36 was initially identified in Japanese and Spanish families as a new type of SCA, characterized by late-onset, slowly progressive cerebellar syndrome often involving motor neurons, or accompanied by sensory neural hearing loss, and cognitive, and emotional disturbances [7, 8]. It is caused by a heterozygous expansion of an intronic GGCCTG hexanucleotide repeat in the NOP56 gene on chromosome 20p13. Unaffected individuals carry 3 to 14 repeats, whereas affected individuals have 650 to 2,500 repeats (OMIM #3614153). However, the range was defined based on only one single research [7] under an obsolete technology and unsuitable statistics, making individuals under the uncertain range for SCA36 diagnosis between 14 to 650 repeats inaccessible to PGT-M.

We recommend an update of knowledge. This study, taking CA36 as an example, aims to integrate updated data, 95% CI under Bayesian methods and long-read sequencing (LRS) to present an up-to-date pathogenic range of SCA36, along with the validation of a specific clinical case, to propose an efficient novel methodology to recalibrate pathogenic repeat expansions.

## Methods

### Participants

The study subjects were members of one generation of a Han Chinese ataxia family pedigree in Sichuan province. A detailed medical history and physical examination record of the proband (II-2) and other family members were evaluated by our multidisciplinary teams including a geneticist, neurologist, otorhinolaryngologist, obstetrician, and gynecologist, reproductive endocrinologist, members of the ethical committee. The blood samples were obtained from participants who asked for carrier screening for SCA36 at the Center for Medical Genetics, Sichuan Academy of Medical Sciences & Sichuan Provincial People’s Hospital. This study was in agreement with the Guidance of the Ministry of Science and Technology for the Review and Approval of Human Genetic Resources. All study procedures were approved by the Ethical Committee of Sichuan Academy of Medical Sciences & Sichuan Provincial People’s Hospital in accordance with the Helsinki Declaration of 1975, as revised in 2000, and patients signed an informed consent form.

### Repeat-primed PCR

Given the initial suspicion of SCA36, repeat-primed polymerase chain reaction (RP-PCR) screening combined with Southern blotting with capillary electrophoresis was employed to identify the presence of SCA36 in the proband and other family members. RP-PCR utilizes repeat primers that amplify alleles, producing PCR products that anneal and form a “ladder” pattern upon separation by capillary electrophoresis. For DNA fragment analysis, the RP-PCR products were processed using the ABI-Prism 3730XL Genetic Analyzer, and the resulting data were analyzed with GeneMarker software.

### Targeted gene capture and PacBio long-read Sequencing

Using a g-tube tube (Covaris, Australia) and centrifugation at 6000 × g, 3 μg of genomic DNA was physically sheared into 8-9 kb fragments after the purity, followed by the evaluation of the integrity of the DNA. The DNA fragments were then connected with barcodes after being repaired by the Damage Repair End Preparation Mix (Vazyme, China). Subsequently, Rapid DNA ligase (Vazyme, China) was used to attach adapters to the DNA fragments. After purifying the ligation products with Agencourt AMPure XP beads (Beckman Coulter, USA), amplification was carried out. The amplified products were captured by hybridization with biotinylated probes (Boke Bioscience, China) that targeted genes relevant to repeat expansion diseases. The SMRTbell Express Template Kit 2.0 (PacBio, USA) was utilized to generate an SMRTbell library. The constructed DNA library was then subjected to long-read sequencing on the PacBio Sequel IIe platform.

After the sequencing data was evaluated and qualified by SMRT Link, bioinformatics analysis of the raw sequencing data was carried out. The particular processes are as follows. The raw sequencing data was analyzed using the official PacBio software package SMRT Link (version 12.0) to generate HiFi reads, and CCS (Circular Consensus Sequence) reads were also automatically generated by the PacBio SMRT Analysis Module, which is an apart of SMRT Link. The data of each sample was divided according to the barcode using the Lima software (version 2.7.1). The CCS reads were aligned to the reference genome GRCh37/hg19 using the Minimap2 software (version 2.24), and the number of repeat units of the gene was calculated using GrandSTR, while the number of repeat units on each read was manually checked using the IGV software.

### Statistical analysis

Considering unaffected individuals typically carry 3 to 14 repeats, whereas affected individuals carry 650 to 2,500 repeats, the pathogenic repeats exhibit a pronounced skewed distribution with one tail. Gamma distribution was hence adopted to describe the pathogenic repeat range. Based on this assumption, we first estimated the prior distribution based on the dataset from [7].

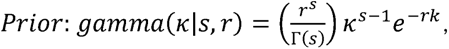

where *k* is a shape parameter, *s* is a scale parameter and *r*, called a rate parameter, is an inverse scale parameter *r = 1/s. k* and *s* are calculated based on the mean and variance of the old data.

Bayesian approaches were applied to update this posterior probability, based on the prior probability and new data. Given two events *A* and *B*, the conditional probability of *A* given that *B* is true is expressed as follows:

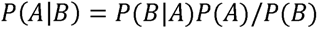

where *P*(*A*) is the prior probability of A to present the experience, *P*(*B*|*A*) is the likelihood function to present the new data, *P*(*A*|*B*) is posterior probability to present the current knowledge, and *P*(*B*) is the total probability of the probability of B with *P*(*B*) ≠ 0. As *P*(*B*) does not change in the same analysis, the formula can be also interpreted as:

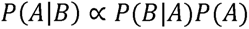

where the current knowledge is proportional to the product of the likelihood function and the new data under the same data distribution. Assume that the new observed dataset consists of *n* observations, denoted as *x*_1_, *x*_2, …_*X*_n_ . Assuming a conjugate prior of the same distribution, the posterior distribution also follows a gamma distribution with updated parameters:

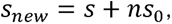

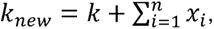

Where *s*_0_ is estimated by the likelihood from the new data [9]. The posterior distribution is then given by:

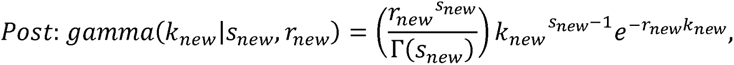

where *r_new_* = 1/*s*_new_. Using this distribution, we calculated a 95% confidence interval (CI) to estimate the range within which the true population parameter is expected to fall with high confidence. This range also defines the new cutoff for normal observations, classifying patients with repeats falling within this range as having the presence of SCA36. Data were analyzed and visualized with R statistical software (v4.2.1; R Core Team 2023)[10].

## Results

### 1 Literature review and summary

We conducted a comprehensive search with the keywords “SCA36” or “Spinocerebellar ataxia-36” across several databases, including PubMed, Embase, Web of Science, and additional platforms such as Dimensions and Semantic Scholar in May 2024, yielding a total of 67 articles. The subsequent filter process is illustrated in Figure 1. To elaborate, we first refined our search publication date from 2012 onwards to include literature published after the research from which the pathogenic range of SCA36 was defined, retaining 55 records. We then meticulously reviewed the abstracts of these articles, excluding 9 duplicates, 11 reviews, 18 basic medical research, 2 case reports, 7 comments, and 2 non-biomedical articles. The remaining 9 articles encompassed detailed data and processing methodologies. Accordingly, Table 1 summarizes these 9 articles, detailing the authors, publication year, ethnic group, sample size, repeat range, mean age at onset, detection method, and references. Upon a thorough examination of the original data from these 9 articles [7, 9, 11–17], we noticed issues such as insufficient data [11–13], questionable data reliability [14], suboptimal data distribution [11], and poor data accessibility [15, 16], and non-Caucasian ethical population [11–14]. Consequently, we determined that only two articles provided data suitable for subsequent analysis [7, 9].

**Figure 1.**
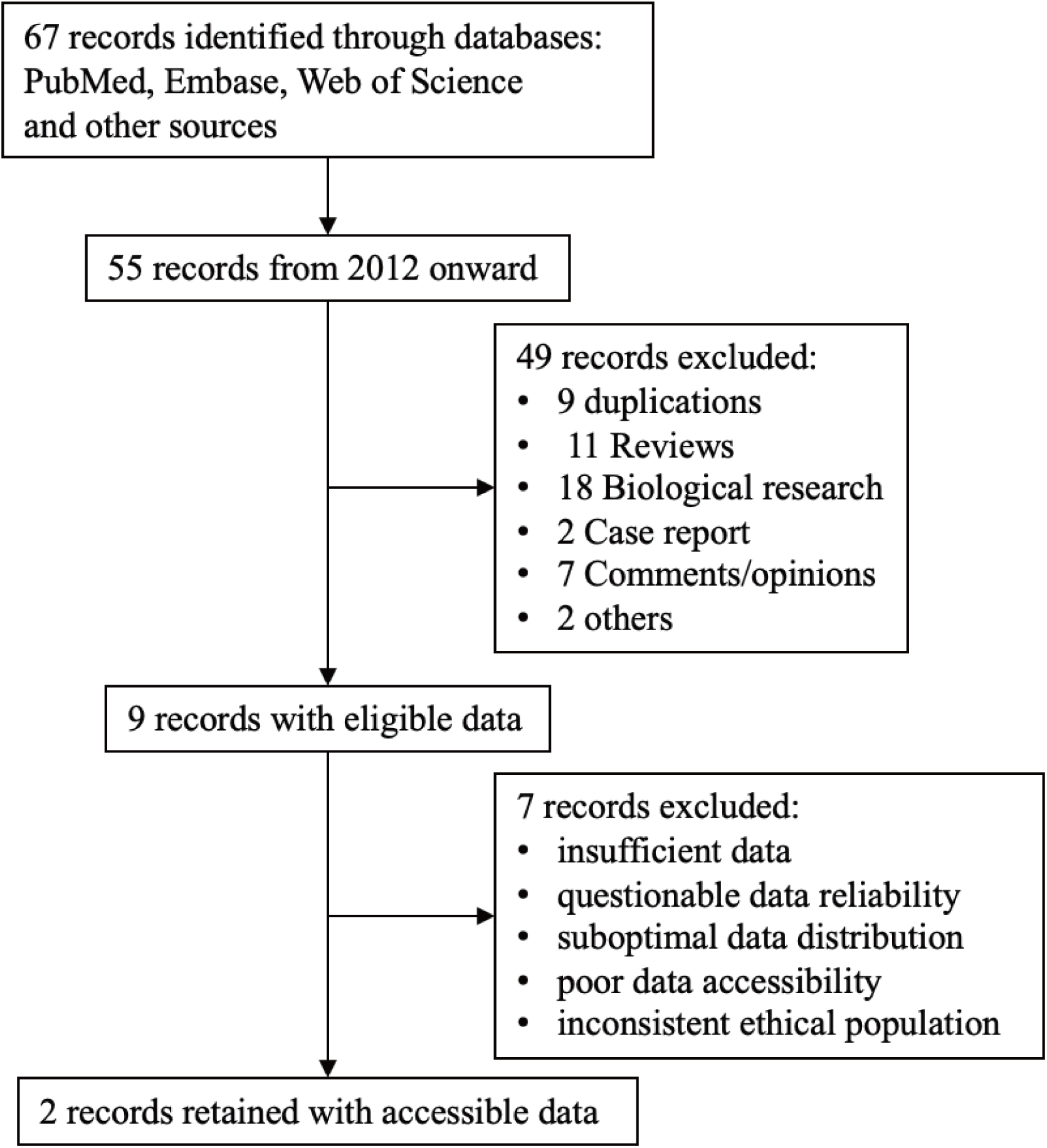
Schematic process of literature review and screening.

**Table 1.** Summary of 9 records with eligible data.

| tim | Year | Ethnic group | Count | Pathogenic repeats | Mean age at onset (years) | Detection method | Reference |
| --- | --- | --- | --- | --- | --- | --- | --- |
| Garcia-Murias et al. | 2012 | Spain | 27 | 650-2500 | $52.5 \pm 7.2$ | Southern blot | [7] |
| Obayashi et al. | 2014 | French/<br>German/Japanese | 17 | 21-2000 | $50.4 \pm 7.2$ | PCR-fragment analyses | [9] |
| Zou et al. | 2023 | Chinese | 3 | 60-2023 | $44.5 \pm 3.8$ | Long-read sequencing | [11] |
| Wang et al. | 2022 | Chinese | 4 | 782 ~ | $45 \pm /$ | Long-read sequencing | [12] |
| Lee et al. | 2016 | Chinese | 11 | Not detected | $44.8 \pm 3.8$ | PCR-fragment analysis | [13] |
| Ikeda et al. | 2012 | Japanese | 18 | 1700-2300 | $52.8 \pm 4.3$ | Southern blot | [14] |
| Lam et al. | 2022 | British | 7 | 30~ | $48.4 \pm /$ | Whole Genome Sequence | [15] |
| Valera et al. | 2017 | USA | 6 | 1500~ | $44.5 \pm 5.8$ | PCR-fragment analyses | [16] |
| Zeng et al. | 2015 | Chinese | 14 | 1000~ | $50.82 \pm 5.02$ | Southern blot | [17] |

### 2 Recalibrating pathogenic range of SCA36 repeat expansions

According to the data published in 2012 by Garcia-Murias et al [7], which Online Mendelian Inheritance in Man (OMIM, https://www.omim.org/) used as a guide, the range of pathogenic repeat expansions is 650 to 2,500. By examining the original data, we found it skewed and inconsistent with the normal distribution. Taking the one-tailed and skewed distribution of the data into consideration, we employed the gamma distribution to simulate the prior distribution, obtaining a 95% CI of [446, +∞]. Since the prior distribution data shares a common original disease with the data published in 2014 by Obayashi et al.[9] we defined them as conjugate distributions. Therefore, we simulated the likelihood function, obtaining a 95% CI of [124, +∞]. Using Bayesian methods, we inferred the posterior distribution and its 95% CI of [484, +∞]. The fitted distribution curves and the integrated curve are depicted in Figure 2. The results indicate that the lower bound of the pathogenic repeat expansions in the posterior distribution is 484, which is less than the 650 as defined in the OMIM guidelines.

**Figure 2.**
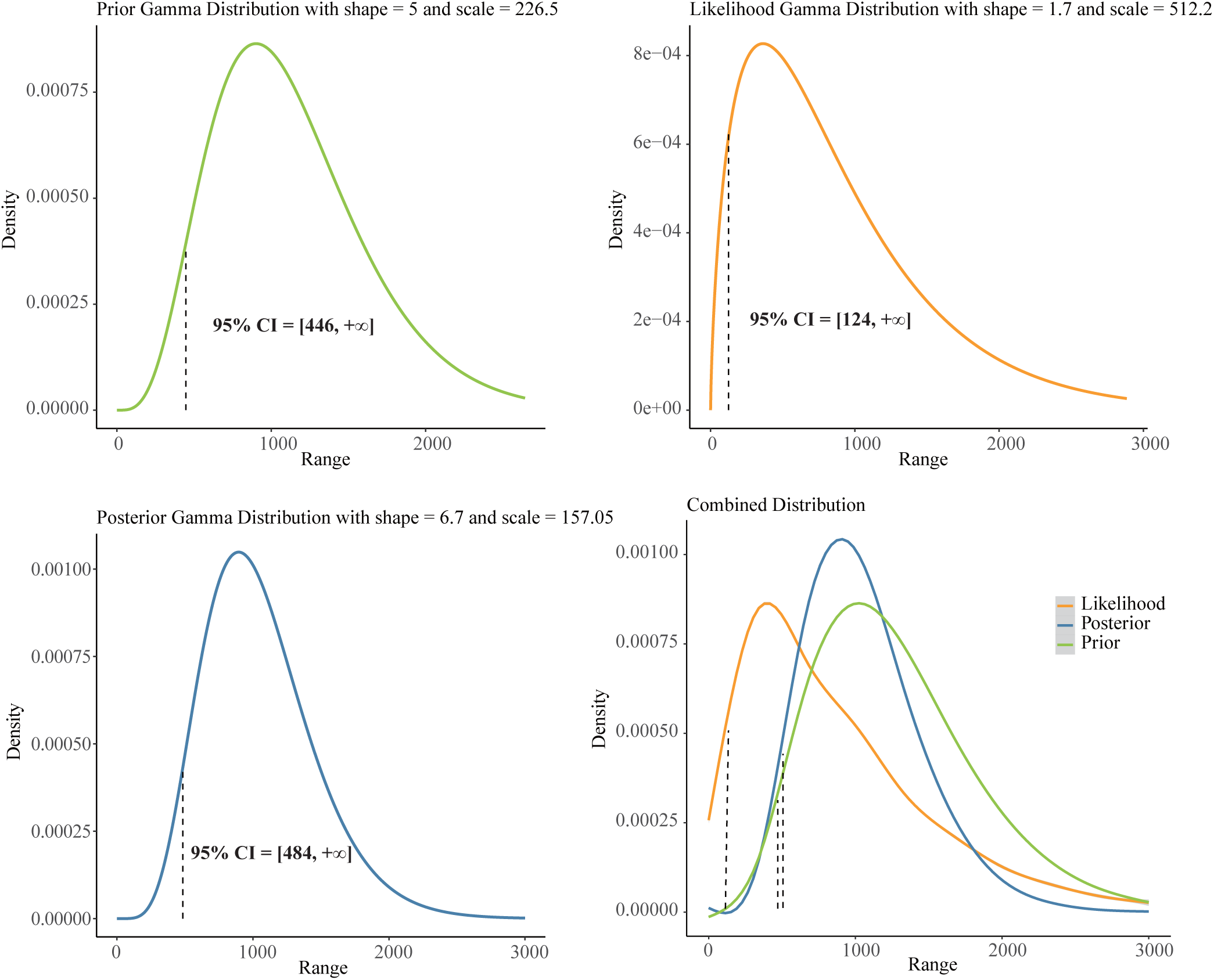
Prior, likelihood, posterior distributions and corresponding 95% confidence interval.

### 3 Case validation

In this study, the proband (II-2) initially presented with progressive symptoms in her 50s, including gait instability, frequent falls, difficulty in lifting the feet, swaying while walking, inability to walk in a straight line, slurred speech, choking on water, dizziness, and slight difficulty in writing. Magnetic resonance imaging indicated cerebellar atrophy (Figure 3B). She did not exhibit bradykinesia, hand tremors, or other Parkinsonian symptoms, and her pain, light touch, and proprioception were intact. Additionally, electromyography did not show significant reductions or the presence of large motor unit potentials. Genetic panel results were negative for SCA1, SCA2, SCA3, SCA6, SCA7, SCA8, SCA12, SCA17, and DRPLA, but revealed that the SCA36 repeats exceeded 14 (Figure 3B). Her daughter (III-1), seeking consultation for assisted reproductive technology, also underwent genetic panel testing, which similarly indicated that the SCA36 repeats exceeded 14 (Supplementary Table_S1). Testing of I-2 was negative, while I-1 was absent due to passing away. Given III-1’s request for PGT-M, we performed long-read sequencing (LRS) to ascertain the precise repeats. Figure 3B illustrates the span of long reads in LRS for II-2 and III-1. Results revealed that II-2 had 418 repeats, while III-1 had 499 repeats, with other potential disease-related gene mutations excluded (Supplementary Table_S2&S3). These findings demonstrate that III-1’s repeats fall within the previously mentioned 95% CI of posterior distribution [484, +∞], thus confirming that III-1 meets the criteria for a positive SCA36 diagnosis within the updated pathogenic range of repeat expansion.

**Figure 3.**
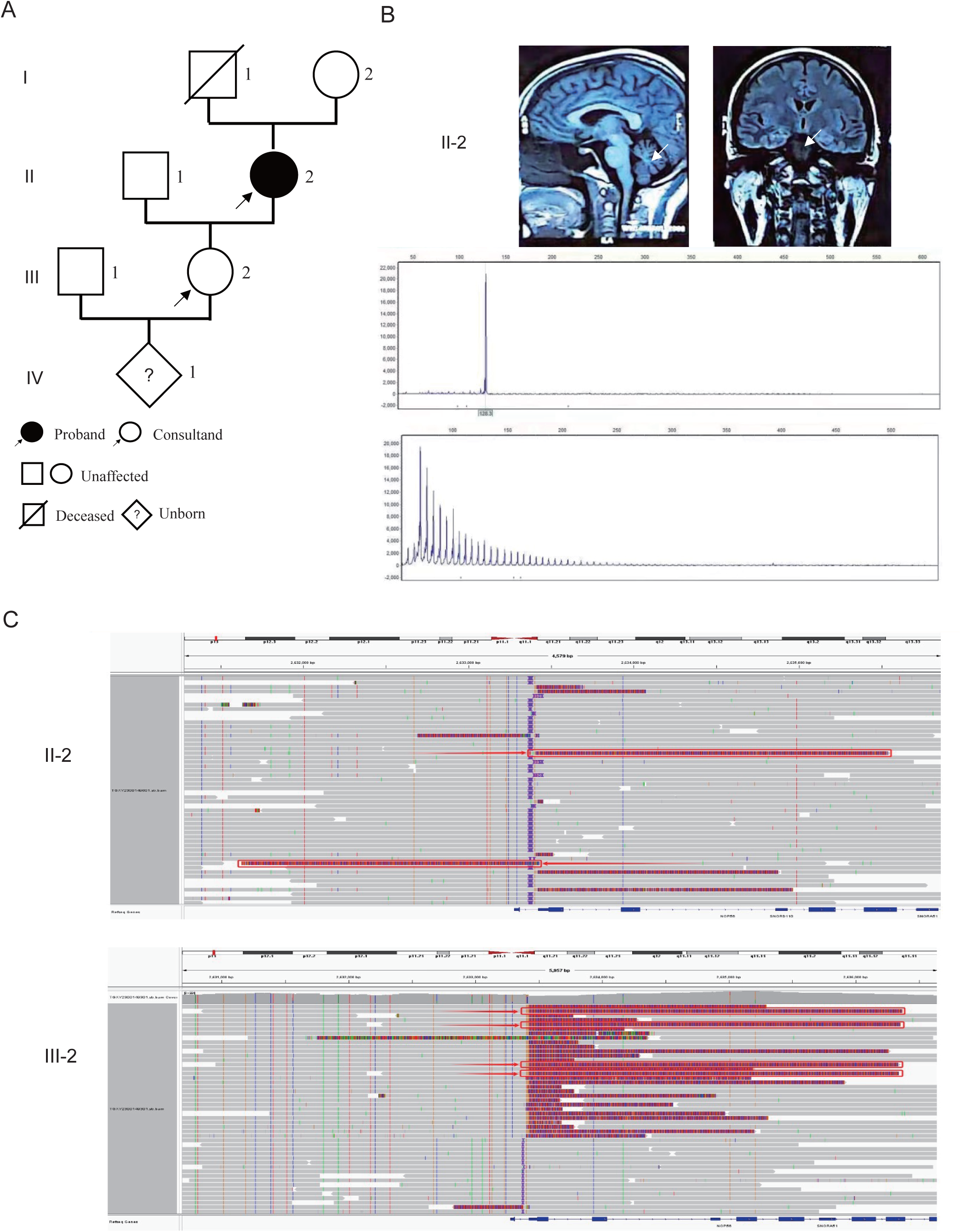
Identification of Expanded GGCCTG Repeat within NOP56 in the SCA family pedigree. (A) The pedigree of a family with spinocerebellar ataxia 36. (B) Magnetic resonance imaging indicates cerebellar atrophy (white arrows) of II-2 and her corresponding conventional PCR shows one peak in an allele and another a characteristic ladder pattern with a typical 6-bp periodicity repeat expansion in NOP56. (C) Long-read sequencing to ascertain the precise repeat range of II-2 and III-1.

## Discussion

The ethical standards for PGT-M in SCA36 patients are stringently ruled and confined to a certain pathogenic range. The overcautious criterion has prevented those SCA36 patients within an uncertain range from performing the PGT-M. In this study, we propose a more inclusive and pragmatic diagnostic methodology to encompass more SCA36 patients in the uncertain range. Initially, we conducted a comprehensive literature review, identifying three original studies with accessible data. Subsequently, we employed Bayesian methods to infer the posterior distribution, obtaining a 95% CI for the distribution. Our results indicated that the lower bound fell below the defined pathogenic range. Lastly, we utilized the LRS technology in the clinical practice to identify the pathogenic repeat expansions in a case, whose repeat expansions also fell within the aforementioned 95% CI, thereby validating the necessity and efficacy of recalibration for repeat expansions.

SCA36 exemplifies cases within an uncertain range that encounter the ambiguity of mutations and phenotypes, displaying an uncertain diagnostic range of 14-650 repeat expansions, within which patients may not exhibit pronounced symptoms, particularly those of childbearing age who have not reached the average onset age of 51.23 ± 7.33 years (male = 51.23 years, female = 51 years) [11]. In our case, III-1 is in her 30s, suggesting that she is likely far from the average age of disease onset. However, SCA36, as a dominant repeat expansion disorder, is characterized by genetic anticipation, where clinical manifestations appear earlier and/or become more severe in successive generations. Dominant repeat expansion disorders have a well-established correlation between expansion size and both age of onset and disease severity, with larger expansions associated with earlier onset and more severe phenotypes [18]. Compared to II-2’s 418 repeats, that III-1 has 499 repeats suggests III-1 might experience earlier onset and more severe symptoms than II-2. To prevent the transmission and exacerbation of the serious condition to her children, the application of PGT-M in assisted reproductive technology is highly warranted.

While the stringent rules for PGT-M in assisted reproductive technology are understandable—primarily aimed at avoiding potential off-targets, especially for monogenic diseases of unknown etiology—such stringent criteria may not be appropriate for the uncertain definition of dynamic mutations. Taking SCA36 into consideration, PGT-M is only permissible when repeat expansions exceed 650. Nevertheless, in our case, both II-2 (with typical manifestations) and III-1 fall below the range. Therefore, it is reasonable to suggest that the defined pathogenic range should be extended more rationally to enhance the application of PGT-M in clinical practice. In this study, the Bayesian methods, along with 95% CI were applied to recalibrate the pathogenic range. Bayesian theorem allows for the integration of prior experience with updated data, produced by new research or novel technologies, to refresh the current knowledge [19]. Thus, an effective method to integrate past and future data is highly beneficial for updating clinical diagnostic guidelines. In our case, the defined pathogenic range of 650 to 2,500 was based on the single research from [7], but with our recalibration based on Bayesian methods the updated range is [484, +∞]. III-1’s repeat expansion therefore falls in, meeting the ethical requirements for PGT-M in assisted reproductive technology. Indeed, we have also conducted a direct integration of the datasets from the two aforementioned studies, yielding a 95% CI of [337, +∞] (Supplementary Figure_S1), which agreed with the conclusion but might undertake the potential overshadowing of information and weight bias from smaller datasets by those with larger sample sizes. Therefore, we advocate for the application of Bayesian methods, which amalgamates prior and likelihood function data, to refine and enhance the integral understanding.

It is important to note that SCA36 is characterized by a heterozygous expansion of an intronic GGCCTG hexanucleotide repeat in the NOP56 gene, resulting in high GC content and extremely long repeat motifs in the repeat expansion [7, 8]. Conventional detection methods are limited. In our case, the patient initially underwent RP-PCR combined with Southern blotting following capillary electrophoresis to measure the GGCCTG repeat expansions, but specific repeat sequences were not obtained. As a matter of fact, over the past decades, the repetitive nature and abundance of short TRs in the human genome have posed significant challenges for genome-wide studies [18]. Short-read sequencing alone fails to accurately map long repeat sequences, leading to covered reads often mapping to multiple genomic regions, thereby being truncated or discarded. Recently, the LRS has emerged in clinical practice to perform a more precise detection of TR expansions [20]. The LRS can obtain reads exceeding 15 kb and sometimes up to 2 Mb, enabling the detection of thousands of structural variants, including repeat regions, in individuals [21]. In this study, compared to RP-PCR combined with Southern blotting, LRS was used for the accurate detection of dynamic mutations to determine whether the repeat expansions exceeded 14 by considering the high GC content and long length. The LRS technology, with its long-read length, high accuracy, and lack of GC bias, provided more specific repeats, thereby offering crucial assistance in determining whether the repeat expansion falls within the recalibrated pathogenic range.

This study has limitations. Firstly, the sample size is limited. We obtained merely 44 patients (27 from the prior and 17 from the likelihood) to recalibrate the pathogenic range. The limited sample size might lead to bias in estimating the true distribution of the data, despite of our attempt to request data from other corresponding authors. Notably, in these limited samples, low repeats between 20 and 30 occurred only in three individuals, casting doubt on whether low repeats were outliers. Fortunately, subsequent cases in Chinese and British cohorts [11, 15] with low repeats reaching 30 or 60 provided proofs to dispute the doubt of the previous data that repeats between 20 to 30 were probably outliers. The second limitation pertains to ethnic differences. Caucasian participants were included in our work to describe the prior, likelihood, and posterior distributions. However, a Han Chinese pedigree as the clinical practice was adopted to validate the recalibration, which might raise the overestimation or underestimation of the pathogenic range given the diversity of gene variations in different ethical populations [22]. This underscores the necessity for further follow-up studies in Caucasian populations, or alternatively, a retrospective analysis with East Asian population to reassess the conclusions.

## Conclusions

To sum up, we proposed a novel methodology that integrates updated data, 95% CI using Bayesian methods and LRS for accurate detection of repeat expansions of dynamic mutations to present an up-to-date pathogenic range of SCA36. We sincerely hope this work would well serve as a strong evidence for those suffering from an uncertain range of repeat expansion.

## Ethics approval and consent to participate

This study was in agreement with the Guidance of the Ministry of Science and Technology for the Review and Approval of Human Genetic Resources. All study procedures were approved by the Ethical Committee of Sichuan Academy of Medical Sciences & Sichuan Provincial People’s Hospital in accordance with the Helsinki Declaration of 1975, as revised in 2000, and patients signed an informed consent form.

## Consent for publication

Patients signed a consent form for publication.

## Availability of data and materials

All data are incorporated into the article and its online supplementary material.

## Data Availability

All data produced in the present work are contained in the manuscript.

## Authors’ contributions

Fulin Liu.: designed the study and wrote the first draft of the manuscript. Wen Huang: participated in the manuscript organization. Ling Liao: supervised the study and edited the first draft of the manuscript. Jiyun Yang: conceived the original idea, and edited the final version of the manuscript.

## Acknowledgements

We thank Beijing GrandOmics Biosciences Co., Ltd. for the technical support on PacBio SMRT target sequencing.

