## Supplementary material for "A Novel Methodology to Recalibrate Pathogenic Range of SCA36 Repeat Expansions for PGT-M": Table_S1_III2.docx

Table_S1 Repeat expansions of common spinocerebellar ataxia-related genes in the Chinese population using high-resolution capillary electrophoresis.

| Disease | Gene | Tandem Repeat | Count |
| --- | --- | --- | --- |
| SCA1 | ATXN1 | (CAG)n | ≤35 |
| SCA2 | ATXN2 | (CAG)n | ≤31 |
| SCA3 | ATXN3 | (CAG)n | ≤44 |
| SCA6 | CACNA1A | (CAG)n | ≤18 |
| SCA7 | ATXN7 | (CAG)n | ≤18 |
| SCA8 | ATXN8/ATXN8OS | (CTA·TAG)n(CTG·CAG)n | <50 |
| SCA12 | PPP2R2B | (CAG)n | ≤23 |
| SCA17 | TBP | (CAG/CAA)n | ≤40 |
| DRPLA | ATN1 | (CAG)n | ≤20 |
