## Supplementary material for "A Novel Methodology to Recalibrate Pathogenic Range of SCA36 Repeat Expansions for PGT-M": Table_S3_III2.pdf

| Individual | NOP56 amplified region_BAM |
|---|---|
| III−2 | 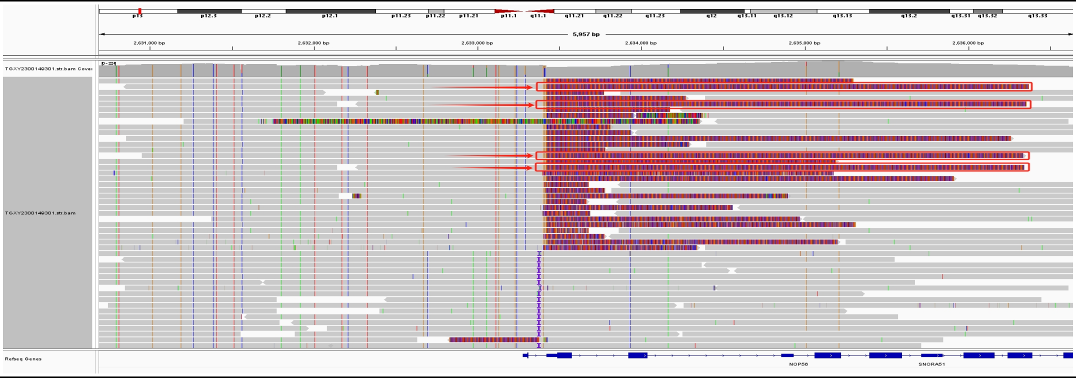 |

Others for detection:

| Dynamic Mutations | | | | | | |
|---|---|---|---|---|---|---|
| Disease | Gene | Nucleotide | Allele count | | Normal | Pathogenic |
| SCA1 | *ATXN1* | CAG | 29 | 30 | 6~35 | ≥39 |
| SCA2 | *ATXN2* | CAG | 22 | 22 | ≤ 31 | ≥ 34 |
| SCA3 | *ATXN3* | CAG | 14 | 32 | 12~44 | 60~87 |
| SCA6 | *CACNA1A* | CAG | 12 | 13 | ≤ 18 | 20~33 |
| SCA7 | *ATXN7* | CAG | 11 | 12 | 7~27 | 37~460 |
| SCA8 | *ATXN8(hg38)/ATXN8OS* | CAG/CTG,CTA/TAG | 9 | 9 | 15~50 | 54~250 |
| SCA10 | *ATXN10* | ATTCT | 15 | 16 | 10~32 | ≥ 800 |
| SCA12 | *PPP2R2B* | CAG | 14 | 16 | 7~31 | 51~78 |
| SCA17 | *TBP* | CAG | 35 | 36 | 25~40 | ≥ 49 |
| SCA27B | *FGF14* | GAA | 9 | 17 | ≤ 250 | ≥ 300 |
| SCA31 | *BEAN1* | TGGAA | 0 | 0 | 14 | 500~760 |
| SCA36 | *NOP56* | GGCCTG | 9 | 499 | 3~14 | ≥ 650 |
| SCA37 | *DAB1* | ATTTT/ATTTC | 9 | 12 | 15 | 31~75 |
| FRDA | *FXN* | GAA | 9 | 9 | 5~33 | ≥ 66 |
| DRPLA | *ATN1* | CAG | 11 | 21 | 6~35 | ≥ 48 |
| CANVAS/MSA | *RFC1* | AAGGG<br>AAAGGG/AAAGG | 11 | 93 | 0~99 | ≥ 100 |
| SCA? | *THAP11* | CAG | 29 | 29 | 20~38 | 45~100 |

| | | | | | | |
|---|---|---|---|---|---|---|
| GDD | *GLS* | GCA | 8 | 13 | 5~38 | 680~1500 |
| FAME1 | *SAMD12* | TTTCA | 0 | 0 | 21 | ≥ 105 |
| FAME2 | *STARD7* | ATTTC | 0 | 0 | 12 | 12 |
| FAME3 | *MARCHF6* | TTTCA | 0 | 0 | 9~20 | 791~1035 |
| FAME4 | *YEATS2* | TTTCA | 0 | 0 | 12 | 192 |
| FAME6 | *TNRC6A* | TTTCA | 0 | 0 | 18 | 29 |
| FAME7 | *RAPGEF2* | TTTCA | 0 | 0 | 18 | 18 |
| SBMA | *AR* | CAG | 21 | 22 | ≤ 34 | ≥ 38 |
| DM1 | *DMPK* | CTG | 13 | 13 | 5~34 | ≥ 50 |
| DM2 | *CNBP* | CCTG | 15 | 16 | ≤ 26 | ≥ 75 |
| OPMD | *PABPN1* | GCN | 10 | 10 | 10 | 11~18 |
| HMN | *VWA1* | GGCGCGGAGC | 2 | 2 | 2 | 1 or 3 |
| ALS | *NIPA1* | GCN | 11 | 12 | 8 | ≥ 10000 |
| AD | *POLG* | CTG | 17 | 18 | ≤ 15 | 10000 |
| HD | *HTT* | CAG | 17 | 18 | ≤ 26 | ≥ 40 |
| HD1 | *PRNP* | CCTCATGGTG GTGGCTGGGG GCAG | 4 | 4 | 4 | 5~16 |
| HD2 | *JPH3* | CTG | 15 | 15 | 6~28 | ≥40 |
| PRS | *EIF4A3* | TCGGCAGCGG CGCAGCGAGG | 7 | 9 | 5~12 | ≥ 15 |
| Fuchs3 | *TCF4* | CTG/CAG | 25 | 29 | ≤ 40 | ≥ 50 |
| Syndactyle1 | *HOXD13* | GCN | 13 | 13 | 15 | ≥ 22 |
| VACTERLX | *ZIC3* | GCN | 9 | 9 | 10 | 12 |
| MRGH | *SOX3* | GCN | 14 | 14 | 15 | 22~26 |

| | | | | | | |
|---|---|---|---|---|---|---|
| BCCD | *RUNX2* | GCN | 16 | 16 | 17 | 20~27 |
| BPES | *FOXL2* | GCN | 14 | 14 | 14 | 15~24 |
| TOF | *TBX1* | GCN | 14 | 14 | 15 | 25 |
| FTDALS1 | *C9orf72* | GGGGCC | 3 | 7 | 2~24 | ≥ 61 |
| NIID/OPDM3 | *NOTCH2NLC* | GGC | 19 | 20 | < 38 | ≥ 66 |
| EPM1A | *CSTB* | CCCCGCCCCGCG | 2 | 3 | 2~3 | ≥30 |
| FXS | *FMR1* | CGG | 27 | 28 | 5~44 | > 200 |
