## Supplementary figures and images for "A Novel Methodology to Recalibrate Pathogenic Range of SCA36 Repeat Expansions for PGT-M"

### Figure_S1.pdf

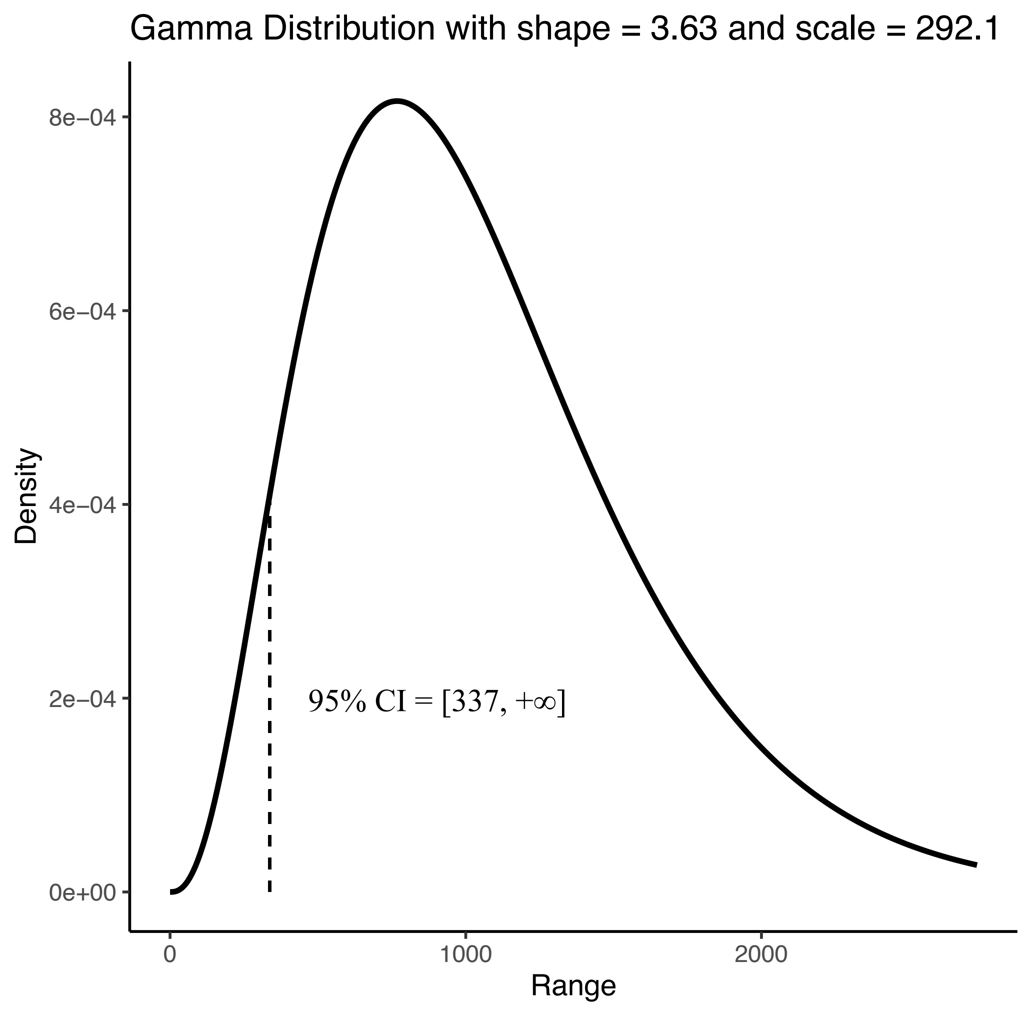
